# Incorporating the Laplacian Filter with a Three-Stream Multi-Channel Convolutional Neural Network for Improved Abnormality Detection in Knee MRIs

**DOI:** 10.1101/2021.01.29.21250504

**Authors:** Rahul Kumar, Rohan Bhansali

## Abstract

Despite ACL and meniscus tears being among the most common movement induced injuries, they are often the most difficult to diagnose due to the variable severity with which these tears occur. Typically, magnetic resonance imaging (MRI) scans are used for diagnosing ligament tears, but performing and analyzing these scans is time consuming and expensive due to the necessitation of a radiologist or professional orthopedic specialist. Consequently, we developed a custom three-stream convolutional neural network (CNN) architecture that contains multiple channels to automate the diagnosis of ACL and meniscus tears from MRI scans. Our algorithm utilizes the sagittal, coronal, and axial slices to maximize feature extraction. Furthermore, we apply the Laplace Operator on the MRI scan images to evaluate and compare its propensity in different medical imaging modalities. The algorithm attained an accuracy of 92.80%, significantly higher than that of orthopedic diagnosis accuracy. Our results point towards the feasibility of shallow, multi-channel CNNs and the ability of the Laplace Operator to improve performance metrics for MRI scan diagnosis.

## 1 Introduction

### 1.1 Magnetic Resonance Imaging in the Knee

Magnetic resonance imaging (MRI) scans are among the most commonly performed diagnostic procedures, with over 40 million performed annually in the United States alone [5]. Performed on the knee, these scans can reveal conditions such as tears in the anterior cruciate ligament (ACL) and meniscus. Specifically, the coronal, sagittal, and axial slices of the MRI are used in conjunction with one another to make the final diagnosis. These three views are shown in Figure 1.The sagittal plane divides the knee into left and right halves and lies on the xz-plane, while the coronal plane divides the knee into front and back and lies on the xy-plane. On the other hand, the axial plane is parallel to the ground, dividing the knee into top and bottom parts. Within the MRI scans, numerous afflictions can be discerned, ranging from damaged cartilage to sprained tendons. More relevantly, MRI scans play an important role in diagnosing ACL and meniscus tears, which often manifest themselves in these scans [6].

**Figure 1:**
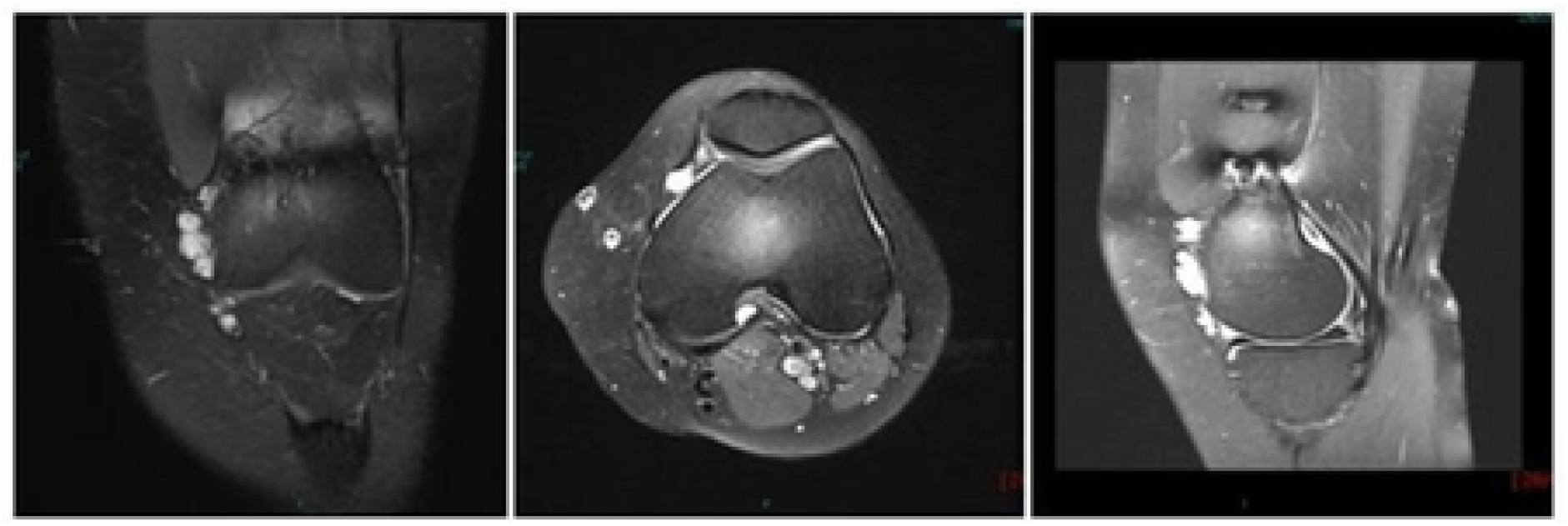
Three views of a knee MRI: coronal, axial, and sagittal respectively.

**Figure 2:**
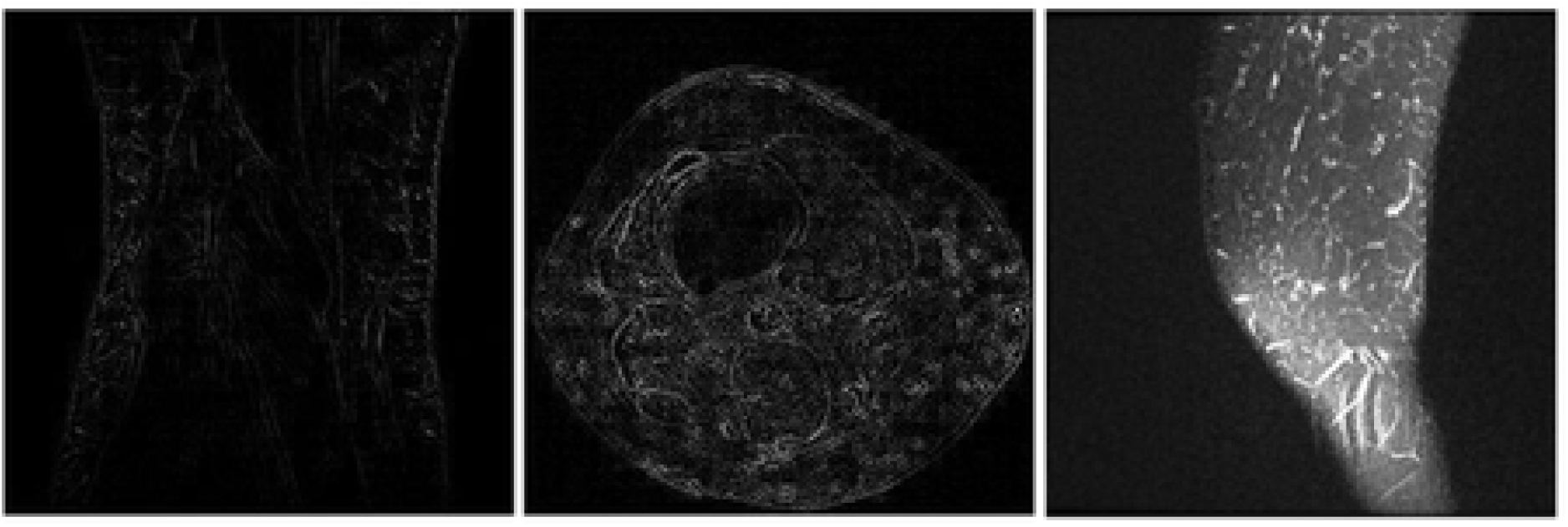
Three views of a knee MRI under the Laplacian Filter: coronal, axial, and sagittal respectively.

### 1.2 Previous Works

Although the field of deep learning in knee MRI diagnosis is still emerging, there has already been some promising research conducted, such as a study recently published by Stanford University [3]. They developed an algorithm called MRNet, capable of detecting abnormalities, ACL tears, and meniscal tears with AUCs of 0.937 (95% CI 0.895, 0.980), 0.965 (95% CI 0.780, 0.914), and 0.847 (95% CI 0.780, 0.914). When compared to radiologists, their model achieved comparable results to radiologists in diagnosing abnormalities but underperformed in detecting ACL and meniscus tears, specifically in the sensitivity metric. Another study provided further evidence for the ability of deep learning models to perform diagnosis tasks at a level of accuracy similar to radiologists [4]. They developed a two-stream convolutional neural network model capable of diagnosing cartilage lesions, including cartilage softening, fibrillation, and fissuring. Their model reached a peak sensitivity of 87.9%, which was well within the confidence interval of several radiologists’ sensitivity score.

### 1.3 Laplace Operator

The Laplace Operator is a second-order differential operator that is used in image processing for accentuating subtleties and otherwise indiscernible properties integral to deep learning classification. It calculates a sum of differences across multiple neighboring pixels to replace the magnitude of each individual pixel and is used specifically to enhance an image’s edges [1]. Previously, we demonstrated the efficacy of the Laplace Operator in the binary classification task of diagnosing COVID-19 from chest CT scans and X-rays with 92% and 94% accuracy, respectively [2]. In this study, we sought to extend the application of the Laplace Operator to the MRI modality while exploring its ability to perform multi-label predictions.

## 2 Methodology

### 2.1 MRNet Dataset

We trained our model on the MRNet dataset published by Stanford University Medical Center containing 1,370 knee MRI images from 1,088 patients [3]. Of these images, 319 (23.3%) were ACL tears, 508 (37.1%) were meniscus tears, and (39.6%) were normal. All MRI exams were performed with GE scanners, with approximately half conducted using a 3.0-T magnetic field and the remaining using a 1.5-T magnetic field. The images were previously scaled to a uniform size of 256×256 pixels.

### 2.2 Training Standards

We created two sets of images, with one set consisting of images after the Laplace Operator was applied and one set consisting of images void of the Laplace Operator. Subsequently, both datasets were split into training and testing sets; 1,124 (90%) images were placed in the former and the remaining 125 (10%) images were placed in the latter. The model was trained with a batch size of ten images over ten epochs using the binary cross-entropy loss function, seen in equation 1. Early stopping was performed after five epochs to prevent over fitting and to increase our model’s ability to generalize on different images.

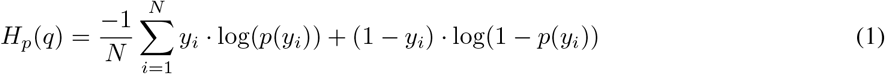

### 2.3 Environment and Device Specifications

The model was developed in Python 3.7.7, using Keras 2.2.5 with a Tensorflow backend.The model was run on an NVIDIA GeForce RTX 2020 with Max-Q Design.

### 2.4 Model Design

We designed a unique architecture that entailed three convolutional neural networks being fused together into a single model. Each of the individual networks had eight layers, with the final layer of each network concatenated together into a combined output layer. The eight layers of the individual networks consisted of an input layer, two convolution layers, a dense layer, two dropout layers, and a flattening layer. After being concatenated together, the fused model contained two more dense layers in addition to another flattening layer. The full model architecture can be seen in Figure 3.

**Figure 3:**
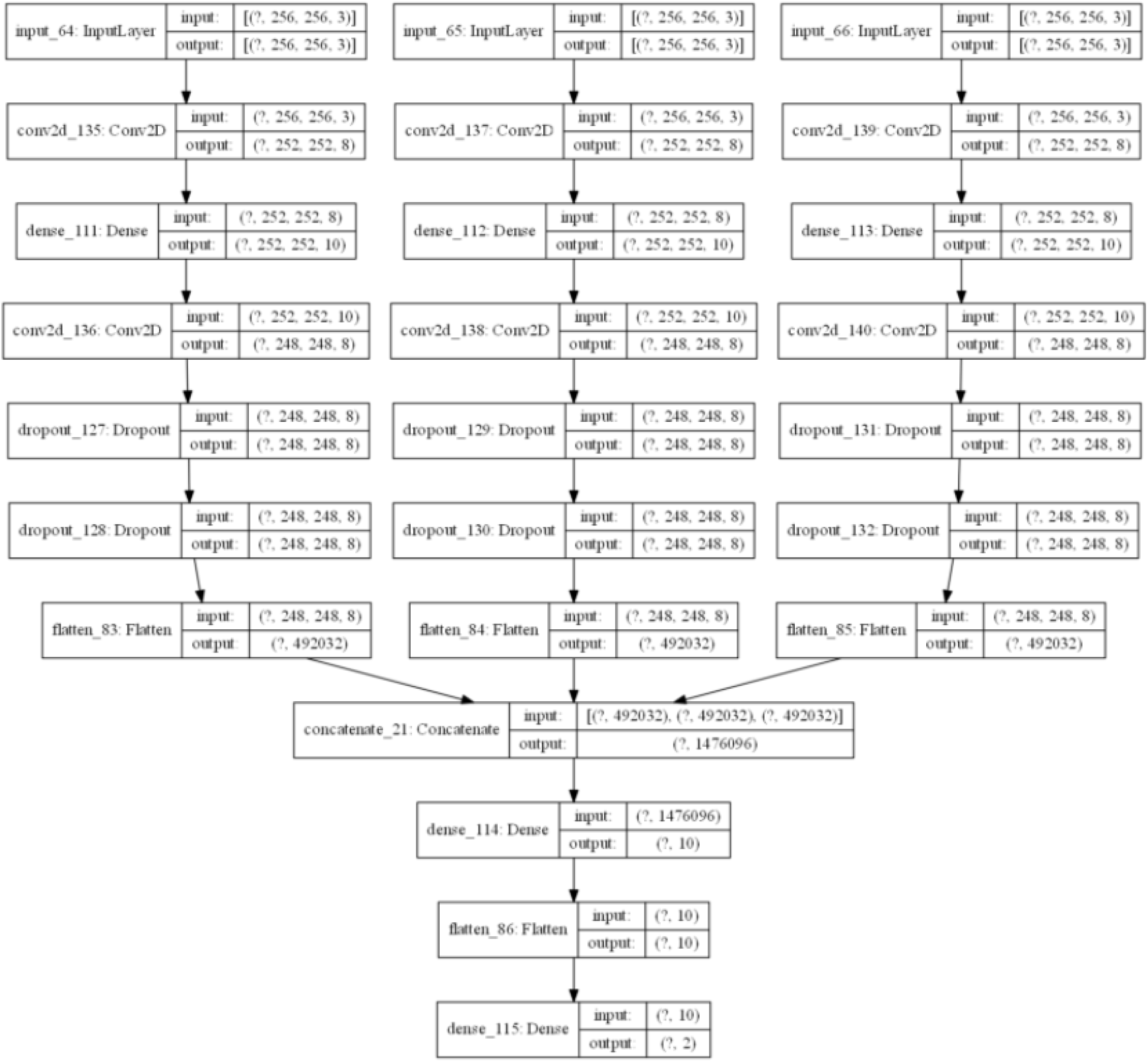
Model Architecture

## 3 Results

The model was trained with a batch size of ten images over ten epochs using the binary cross-entropy loss function. Early stopping was performed after five epochs to prevent overfitting and to increase our model’s ability to generalize on different images. The model achieved an accuracy of 89.2% on the set of images without the Laplace Operator performed and an accuracy of 92.8% on the set of images with the Laplace Operator performed.

## 4 Conclusion

We trained a novel three-stream convolutional neural network model to detect ACL and meniscus tears from knee MRIs using the MRNet dataset, which consisted of 1,249 images provided by researchers at Stanford University. Building upon our past success using the Laplace Operator in medical imaging diagnosis, we performed this filter on the images within our dataset and found that our model attained an accuracy of 92.8%, which equated to higher overall performance as compared to the 89.2% accuracy of our model when trained on images without the Laplace Operator. Therefore, our results provide support for the efficacy of the Laplace Operator in boosting the performance of deep learning models in diagnosing MRI scans. Moreover, we demonstrate the ability of our custom three-stream, multi-channel convolutional neural network in predicting the presence of ACL and meniscus tears.

## Data Availability

This data was obtained from Stanford University School of Medicine. It is available at this url: https://stanfordmlgroup.github.io/competitions/mrnet/. In order to obtain the data, the Research Use Agreement was signed, which is available at the aforementioned link. As part of the Research Use Agreement, the authors consented to this data only being used for non-commercial, research purposes. Adequate credit has been allocated to Stanford University School of Medicine in the paper citations. Stanford University has validated the ethical collection of the data. All subjects in the dataset are anonymized.

https://aimi.stanford.edu/mrnet-knee-mris

## References

[1] Bhansali, R., Kumar, R., Writer, D. CoronaNet: A Novel Deep Learning Model for COVID-19 Detection in CT Scans. In Journal of Student Research 9(2), doi: https://doi.org10.47611/jsrhs.v9i2.1246, JOFSR, 2020.

[2] Bhansali, R., Kumar, R., Writer, D. CoronaNeXt: Evaluating the Performance of the Laplacian Operator in Diagnosing COVID-19 From Chest X-rays. In International Journal of Engineering Research and Technology 9(11), doi: https://dx.doi.org/10.17577/IJERTV9IS110211, JOFSR, 2020.

[3] Bien, N., Rajpurkar, P., Ball, R. L., Irvin, J., Park, A., Jones, E., … Lungren, M. P. Deep-learning-assisted diagnosis for knee magnetic resonance imaging: Development and retrospective validation of MRNet. In PLOS Medicine 15(11), doi: https://doi.org/10.1371/journal.pmed.1002699, 2018.

[4] Recht, M. P., Zbontar, J., Sodickson, D. K., Knoll, F., Yakubova, N., Sriram, A., … Zitnick, C. L. Using Deep Learning to Accelerate Knee MRI at 3 T: Results of an Interchangeability Study. In American Journal of Roentgenology 215(6), pages:1421-1429, doi:https://doi.org/10.2214/ajr.20.23313

[5] Subhas, N., Patel, S. H., Obuchowski, N. A., Jones, M. H. Value of Knee MRI in the Diagnosis and Management of Knee Disorders. In Orthopedics 37(2), doi: https://doi.org/10.3928/01477447-20140124-11

[6] U.S. National Library of Medicine. Knee MRI scan In MedlinePlus Medical Encyclopedia, https://medlineplus.gov/ency/article/007361.htm.

